# Does a new randomized trial change the evidence? A pilot audit of evidence deltas in colorectal surgery

**DOI:** 10.64898/2026.09.04.26362222

**Authors:** Sang-ji Choi, Gibong Chae

**Affiliations:** School of Medicine, Kangwon National University, Chuncheon, Republic of Korea; Department of Surgery, Kangwon National University Hospital, Chuncheon, Republic of Korea

**Author notes:** **Corresponding author:** Sang-ji Choi, M.D., Ph.D., Department of Surgery, Kangwon National University Hospital, 156 Baengnyeong-ro, Chuncheon 24289, Republic of Korea.

**Keywords:** meta-research, cumulative meta-analysis, research waste, evidence synthesis, colorectal surgery, reporting

## Abstract

**Background:** Research output is counted in publications, although publications differ in how much uncertainty they resolve. An evidence delta is the change between the evidence state immediately before an index trial’s results are published and the evidence state after. Whether such a comparison can be reconstructed from the published record is unknown.

**Methods:** Six colorectal-surgery evidence lineages were chosen by the first author before any outcome data were seen, each defined by one binary patient-relevant outcome. Randomized trials of any year were sought in PubMed and PubMed Central with multiple keyword and acronym strategies. One verbatim strategy per lineage was written afterwards, re-run and archived; the records it returned that had not already been assessed were screened on title and publication type, and those that survived were assessed in duplicate. For every index trial published from 2016 through 2025 with at least one exchangeable earlier trial, the synthesis of all trials published before it was compared with the synthesis including it. The outcome was the proportion of index trials that changed a clinical decision category, defined on the risk-ratio scale with a region of no clinically important difference of 0.90 to 1.11. Categories and thresholds were fixed in an analysis plan frozen before extraction, which was not publicly registered; the interval method used in the main analysis was changed after the first run, and the frozen method is reported alongside. Study selection and data extraction were performed independently by two reviewers, with disagreements resolved by discussion and consensus, and every extracted count was verified against the source record.

**Results:** Of 200 records assessed, 93 were eligible randomized trials and 42 reported the lineage outcome in a form recoverable per treatment group, so 41 trials entered five syntheses covering 11,817 analyzed patients. One lineage could not be analyzed at all. Twenty-nine index trials qualified. Depending on the interval method and the decision threshold, 0 to 1 of 29 index trials changed the decision category, and no trial did so in every specification in which it was included; in the amended main analysis the figure was 1 of 29 (3.4%; 95% CI, 0.6 to 17.2). Twenty-two index trials (76%), covering 8,207 analyzed patients, produced neither a category change nor a 20% gain in precision, and four trials reduced precision. Among the 14 index trials with retrievable full text, 8 cited a prior systematic review, 5 used one to justify the trial, and none presented an updated quantitative synthesis including its own result.

**Conclusions:** Evidence deltas were reconstructable, and decision-category changes were rare and sensitive to analytic specification. The larger obstacle was reporting: 55% of eligible randomized trials could not enter such an accounting at all, because the outcome was reported without recoverable per-arm counts, reported only in another form, not measured, or reported with counts and percentages that could not be reconciled. This purposive, single-database pilot with lineage selection by the first author estimates feasibility rather than prevalence.

## Introduction

Research output is counted in publications, and publications are weighted by the journals that carry them. Neither measure describes how much uncertainty a study resolved. A trial that changes practice, a trial that establishes equivalence, and a trial that adds little to an already stable estimate each yield one publication. Avoidable waste in the production and reporting of research was described more than fifteen years ago and remains substantial.[1] Meta-research has since shown that new studies are inconsistently justified by systematic reviews and that their results are rarely placed in the context of an updated synthesis.[2,3] The literature therefore records what was published more reliably than what the publication changed.

Cumulative meta-analysis offers a way to make that change visible. Adding trials in chronological order shows when an effect estimate stabilized, and it has been used to show that some questions were settled years before trials stopped being performed.[4] What has not been established is whether the same reconstruction can be performed prospectively, trial by trial, as a routine accounting of contribution, using only what investigators actually publish.

This study proposes such an accounting, which we call an evidence delta: the difference between the evidence state immediately before an index trial’s results were published and the evidence state after they were, measured on separate dimensions rather than reduced to a single score. The construct is deliberately multidimensional. A trial may change the clinical decision that the evidence supports, narrow the interval around a pooled estimate, alter heterogeneity, add a population or setting, or leave the previous position intact. None of these dimensions is a measure of scientific worth, and a small measured delta is not evidence of waste.

Measuring such a delta requires two things that the published literature does not obviously supply. The first is a defensible grouping of trials that address the same clinical decision, since a synthesis that mixes non-exchangeable populations or outcome definitions will manufacture change where none exists. The second is arithmetic that a reader can reproduce: the number of patients with the outcome and the number analyzed, in each arm, for the prespecified lineage outcome. We are not aware of a quantification of whether trial reports satisfy the second requirement in a surgical field, although comparisons of trial protocols with their publications have shown that a large share of outcomes are reported with too little data for meta-analysis.[7]

Colorectal surgery is a useful test bed. Its questions are pragmatic, its outcomes are mostly binary and patient-relevant, and its trials are numerous enough to form lineages of exchangeable studies. This pilot asks three questions. Can the evidence states immediately before and after an index trial’s results be reconstructed from published reports? How often does adding a trial change the clinical decision the evidence supports? And what fraction of randomized trials can enter such an accounting at all?

## Methods

### Design and registration

This is a pilot meta-research study with cumulative meta-analysis, conducted as a feasibility test of a larger protocol. A statistical analysis plan specifying the sampling frame, the analysis population, the effect measure, the decision categories, the decision thresholds and the material precision threshold was written and frozen before any outcome data were extracted. The plan was completed before outcome extraction, and it was not prospectively registered or publicly time-stamped. The word prespecified is therefore used below only for the elements fixed in that file, and not for the interval method used in the main analysis or for the post-hoc exclusion analyses. The frozen plan, all amendments, the screening list, the extraction dataset, the verification log and the analysis code are deposited (Data availability). Five amendments were made after the analysis plan was frozen and are reported in the Results and in the deposited amendment file.

### Evidence lineages and eligibility

Six lineages were fixed before any outcome data were seen, each defined by one population, one intervention and comparator pair, and one binary patient-relevant outcome: indocyanine green fluorescence angiography versus none for anastomotic leak; oral antibiotic prophylaxis versus none for surgical site infection; prophylactic mesh at permanent end colostomy versus none for parastomal hernia; intracorporeal versus extracorporeal anastomosis for postoperative complications; early versus late closure of a defunctioning ileostomy for postoperative complications; and prehabilitation versus standard care for postoperative complications.

The six lineages were chosen by the first author before any outcome data were extracted, from questions in colorectal surgery that he already knew to contain several randomized trials, on three criteria: a single binary patient-relevant outcome shared across trials, at least three randomized trials of which at least one was published from 2016 through 2025, and at least one earlier trial so that a prior evidence state existed. The third criterion was not met in practice by three of the six lineages, whose earliest trial falls inside the study window; those trials are reported as lineage- origin trials and contribute no index-trial observation, and the deviation is recorded as an amendment. Candidate lineages were not enumerated systematically, no sampling frame of colorectal-surgery questions was constructed, and the first author had general prior awareness of the direction of results in some of these fields. Lineage selection is therefore a potential source of bias, and the six lineages should not be read as representative of colorectal surgery.

Within each lineage, randomized trials of any publication year were sought in PubMed and PubMed Central using multiple keyword and acronym strategies, with recent systematic reviews used as maps to trial names. Every trial that became a candidate for a synthesis was confirmed from its own PubMed record. The records retrieved by that first search were then screened a second time, independently and from the eligibility rules alone, by the second reviewer, who had not seen the decisions of the first pass; this added four trials that the first pass had set aside and removed one that it had wrongly retained, and every discrepancy was resolved by discussion between the two reviewers and confirmed against the record. Those strategies were run interactively and their strings were not archived at the time. To make the search auditable, one verbatim strategy per lineage was written afterwards and run against PubMed on the final search date; the six strings, their yields and the reconciliation are deposited. That verification search returned 667 records, of which 582 had not been assessed. Their titles and publication types were screened, and 101 records were carried to record-level assessment against the full eligibility rules by two reviewers (S.J.C. and G.B.C.) working independently and blinded to each other’s decisions. The two agreed on 97 of the 101, and the remaining four were resolved by discussion between the reviewers and settled by consensus against the records. One previously unassessed eligible trial was found and added. Thirteen records assessed in the original search, four of them trials in the synthesis, are not returned by the documented strategy for their lineage, which measures the gap between the reconstruction and the original process. This remains a pilot-level search of one database, and it is not a systematic three-database search with a peer-reviewed strategy. Records excluded for design, population, comparator or non-retrievability are listed individually with reasons in the deposited screening file.

An index trial was a randomized trial whose primary report was published between January 1, 2016 and December 31, 2025 in an included lineage. The main analysis population comprised index trials with at least one exchangeable earlier trial in the same lineage and with per-arm events and analyzed denominators recoverable from the published report without imputation from a P value.

### Extraction and verification

Data were extracted independently and in duplicate by two reviewers using a standardized extraction form, and disagreements were resolved by discussion and consensus. Each per-arm count was extracted with the verbatim text containing it and its PubMed identifier; for some trials the stored string is a fragment rather than a whole sentence, in which case the automated string check below is not the step that verifies the count, and the deposited audit log says so. Counts that could not be quoted were recorded as unverified and the trial was excluded rather than imputed. Five verification steps followed. First, an automated check required each quoted fragment to appear in the retrieved PubMed record. Second, every count taken from a full text was re-checked against the article itself by the second reviewer, who had to confirm or refute each extracted count against the source text with a supporting quotation. Third, every count obtained arithmetically rather than printed as an integer was audited for uniqueness: whether an integer numerator exists that reproduces the published percentage on the published denominator, and whether it is the only one. Fourth, a consistency screen searched each arm’s implied percentage in the source abstract. Trials whose published counts and percentages could not be reconciled were excluded, and the arithmetic for each exclusion is given in the Results. Fifth, all four counts for every trial in the syntheses were extracted a second time from the sources by the second reviewer, who had no access to the first extraction, and the two datasets were compared cell by cell; the one disagreement was resolved by discussion and is reported in the Results.

### Synthesis and outcomes

All lineage outcomes were undesirable events, so a risk ratio below 1 favors the intervention. Searches were run on 4 September 2026, which is the final search date. Trials were ordered within a lineage by the publication date recorded in the PubMed record of the primary report, which is the date PubMed carries for the article, whether that is an electronic-ahead-of-print date or a journal issue date. Where a record gave no month, mid-year was imputed; this affected four records, each the only trial of its year in its lineage, so no ordering depended on the imputation. No two trials in the same lineage shared a publication year and month, so no tie-breaking rule was needed. For each index trial, the pre-index-result synthesis included all trials in the lineage published before it, and the post-index-result synthesis added the index trial. Because trials were ordered by publication date, this comparison captures the evidence state immediately before the index trial’s results became available, and not the evidence state at the time the trial was designed or begun. Other trials may have been published while the index trial was recruiting. The parent protocol defines a pretrial evidence date as the day before first enrollment; that definition was not implemented here. This study therefore measures achieved evidence deltas only, and it cannot speak to the intended evidence delta at trial initiation. Random-effects inverse-variance models on the log risk ratio were fitted with restricted maximum likelihood. A 0.5 continuity correction was applied to the single trial with a zero cell.

The main outcome was the proportion of index trials that changed the clinical decision category. Decision categories, decision thresholds and the material precision threshold were fixed in the frozen plan; the interval method used in the main analysis was not. Categories were defined from the 95% confidence interval of the pooled risk ratio against a region of no clinically important difference of 0.90 to 1.11: favors intervention, no clinically important difference, favors comparator, or inconclusive when the interval spanned more than one region. Because no single core outcome set covers these six questions, the region was set at a 10% relative change and tested at 5% and 20%.

Secondary outcomes were the precision gain, defined as one minus the ratio of the post-index- result to the pre-index-result interval width on the log scale, with a material gain prespecified at 20% and tested at 10% and 30%; the relative change in the pooled estimate; the proportional increase in inverse-variance information; changes in heterogeneity; and a descriptive contribution class. A contribution was classified as discovery only when the index trial’s own effect fell outside the 95% prediction interval of the pre-index-result synthesis, which is estimable from three or more prior trials.

Contextualization was assessed for index trials with retrievable full text: whether a prior systematic review was cited in the introduction, whether it was used to justify the trial, and whether the discussion placed the result against a prior synthesis. A fourth item, whether an updated quantitative synthesis including the new result was presented, was added after the plan was frozen and is post-hoc.

Sensitivity analyses used Hartung-Knapp-Sidik-Jonkman intervals and their modification for meta-analyses with few studies,[5] DerSimonian-Laird estimation,[8] fixed-effect models, alternative decision regions and precision thresholds, exclusion of trials with arithmetically derived counts, exclusion of three trials whose populations or outcome definitions sit least comfortably in their lineage, and leave-one-lineage-out analysis. Table 2 reports the fourteen specifications that can change a decision category, of which five were prespecified in the frozen plan and nine were added after the first analysis run, and states which is which. The two alternative precision thresholds cannot change a decision category and are reported in the Results. Analyses were performed in Python 3 with numpy and scipy, following published guidance for meta-analysis in Python.[6] Proportions are reported with 95% Wilson intervals.

## Results

### Evaluability of the published record

Two hundred records were assessed at record level across the six lineages, 99 in the original search and 101 more in the verification search. One hundred and seven were excluded for design, population, comparator or non-retrievability, leaving 93 eligible randomized trials. Of the exclusions, 38 are secondary, follow-up, cost, quality-of-life or other companion reports of trials already assessed, 30 report no randomized comparison, 21 have the wrong comparator, 14 the wrong population, 3 are study protocols and 1 was not retrievable. In 42 of these the number of patients with the lineage outcome and the analyzed denominator were recoverable per treatment group and internally consistent (Table 1).

**Table 1.** Evaluability of randomized trials, by evidence lineage. “Eligible” excludes records set aside for design, population, comparator or non-retrievability; per-trial reasons are in the deposited screening file.

| Lineage | Assessed | Eligible | Outcome recoverable per arm | Entered synthesis |
| --- | --- | --- | --- | --- |
| ICG fluorescence angiography vs none (anastomotic leak) | 20 | 7 | 6 | 6 |
| Oral antibiotic prophylaxis vs none (surgical site infection) | 47 | 28 | 17 | 17 |
| Prophylactic mesh at end colostomy vs none (parastomal hernia) | 29 | 14 | 8 | 8 |
| Intracorporeal vs extracorporeal anastomosis (any complication) | 17 | 8 | 1 | 0 |
| Early vs late ileostomy closure (postoperative complications) | 19 | 11 | 4 | 4 |
| Prehabilitation vs standard care (any complication) | 68 | 25 | 6 | 6 |
| **Total** | **200** | **93** | **42** | **41** |

#### Fifty-one of 93 eligible trials (55%) were not evaluable

The reasons fall into four groups: the outcome was reported without recoverable per-arm counts, for example as percentages without denominators or as a bare P value; it was reported only in another form, such as a composite endpoint, a severity index, event counts rather than patients affected, or a single component such as incisional infection; it was not measured; or the reported counts could not be reconciled with the trial’s own published percentages. Three trials that had entered the syntheses fell into the last group and were removed after audit; three further records were set aside for the same reason before extraction. In PILLAR III, no integer numerator over the reported denominator of 169 yields the reported 9.6% (16 of 169 is 9.5%, 17 of 169 is 10.1%). In the trial by Vierimaa and colleagues, 12 control events are reported as 32.3% although 12 of 35 is 34.3%, and other control percentages in the same abstract imply a denominator of 31 or 32. In PARTHENOPE, 9 mesh-arm events are reported as 12.7%, which no denominator at or below the arm size of 55 can produce.

The second independent screening of the original search added four trials to the syntheses, Taylor 1994 and SOAP 2021 and MECCA 2024 to the oral-antibiotic lineage and Vogel 2023 to the ileostomy-closure lineage, and removed one, the trial by Lopez-Rodriguez-Arias and colleagues, which reports a 20-patient subset recruited during the COVID-19 period from a trial whose main report is published separately. The verification search added one further trial, Clarke 1977, to the oral-antibiotic lineage. None of these changes altered which trial changes the decision category in the main analysis; they changed the denominator, so the main-analysis proportion moved from 1 of 27 to 1 of 29, and they narrowed the range across specifications from 0 to 4 of 27 to 0 to 1 of 29, mainly by removing the two opposite-direction transitions that the Hartung-Knapp specifications had produced in the oral-antibiotic lineage.

The lineage comparing intracorporeal with extracorporeal anastomosis had one evaluable trial and could not be analyzed; that trial’s counts were not extracted, because a lineage with one trial has no prior evidence state, so it is counted in Table 1 on the basis of its screening status alone. Five lineages remained, containing 41 trials and 11,817 analyzed patients.

The independent duplicate extraction reproduced 40 of the 41 trials’ four counts exactly. The single disagreement was Lewis 2002, where the second extraction took the denominators after post-randomization exclusion (104 and 103) rather than the randomized denominators (109 and 106). Both pairs give the published point estimate of 0.29 and the published lower limit of 0.11; they differ in the upper limit, which is 0.748 for 5 of 109 versus 17 of 106 and 0.760 for 5 of 104 versus 17 of 103, against a published 0.75. After discussion between the two reviewers the randomized denominators were retained, and no value changed.

### Decision-category change

Twenty-nine index trials published from 2016 through 2025 had at least one exchangeable earlier trial and were evaluable; they analyzed 10,301 patients. Three further trials were the origin of their lineage within the study window and had no prior evidence state.

#### Depending on the interval method and the decision threshold, 0 to 1 of 29 index trials changed the decision category, and no trial did so in every specification in which it was included (Table 2)

**Table 2.**
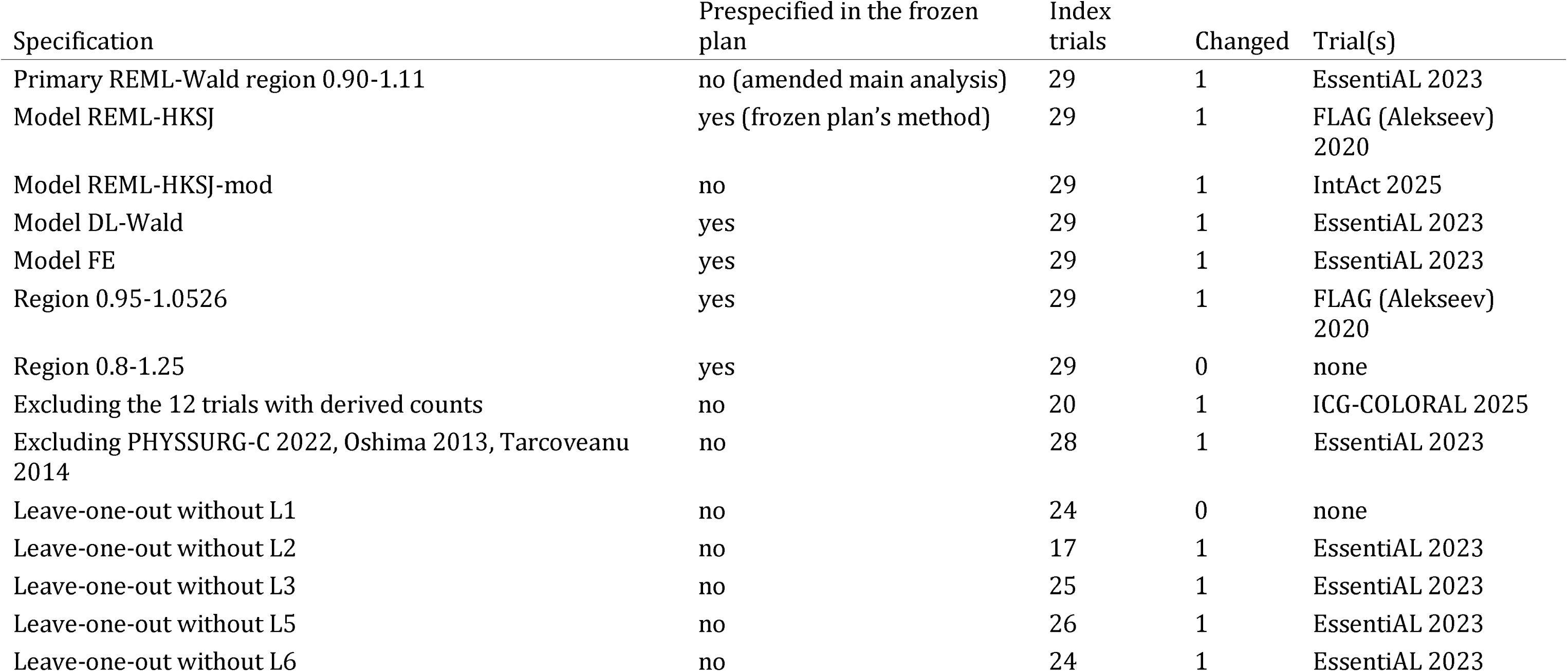
Decision-category changes across the amended main analysis and the sensitivity analyses. Categories: 1 favors intervention, 2 no clinically important difference, 3 favors comparator, 4 inconclusive.

In the post-amendment main analysis the figure was 1 of 29 (3.4%; 95% CI, 0.6 to 17.2). That trial was EssentiAL, which moved the fluorescence-angiography lineage from inconclusive to favors intervention: pooled risk ratio 0.56 (95% CI, 0.35 to 0.90) before and 0.61 (0.44 to 0.83) after. The change was boundary-dependent, since the pre-index-result upper limit was 0.904 against a boundary of 0.90.

Across the 14 specifications examined (Table 2), of which five were prespecified in the frozen plan and nine were added after the first analysis run, the count barely moved but the identity of the changing trial did. Under the frozen plan’s Hartung-Knapp method and under a 5% decision region the changing trial was FLAG rather than EssentiAL, and under the modified Hartung-Knapp method it was IntAct. Under a 20% decision region no trial changed the category. EssentiAL changed the category in 8 of the 12 specifications that included it, FLAG in 2 of 13, and IntAct and ICG-COLORAL in 1 of 13 each. The highest proportion, 1 of 17 (5.9%), arose when the oral- antibiotic lineage was left out, an analysis with a reduced denominator.

### Other dimensions of the evidence delta

Twenty-two of 29 index trials (75.9%), covering 8,207 analyzed patients, produced neither a decision-category change nor a material gain in precision and were classified as low observed delta. Six (20.7%) contributed precision only, and one contributed a category change. No reversal was observed, and no discovery, although the discovery class was unavailable for the 6 index trials whose lineages contained fewer than three prior trials, including the trial that changed the category.

The median precision gain contributed by an index trial was 11.7% (IQR, 5.4 to 19.6), and 7 of 29 (24.1%) reached the 20% threshold, 15 of 29 (51.7%) the 10% threshold and 6 of 29 (20.7%) the 30% threshold. Four trials reduced precision, meaning the pooled interval was wider after their addition than before: Ikeda 2016 (-28.7%), Rybakov 2020 (-1.5%), STOMAMESH (-43.8%) and Berkel 2022 (-23.3%). The median relative change in the pooled risk ratio was 6.6%, and the median increase in inverse-variance information was 19%. Heterogeneity of the post-index-result syntheses had a median I-squared of 32.3% (range 0 to 81.8), and adding the index trial raised I-squared in 6 of 29 syntheses and lowered it in 13.

Lineage trajectories differed (Figure 1, Table 3). In the oral-antibiotic lineage the pooled estimate already favored prophylaxis before the study window (risk ratio 0.38; 95% CI, 0.27 to 0.53 after five trials), and twelve subsequent index trials analyzing 4,737 patients left the category unchanged, ending at 0.46 (0.38 to 0.55). In the prophylactic-mesh lineage the category was likewise fixed before the window and unchanged after four index trials, although STOMAMESH, the largest trial in that synthesis, reported no difference and widened the pooled interval by 43.8%. The early-closure and prehabilitation lineages remained inconclusive throughout, with the prehabilitation estimate moving from 3.30 (0.85 to 12.75) based on a single trial of 21 patients to 0.98 (0.71 to 1.34) after five further index trials.

**Figure 1.**
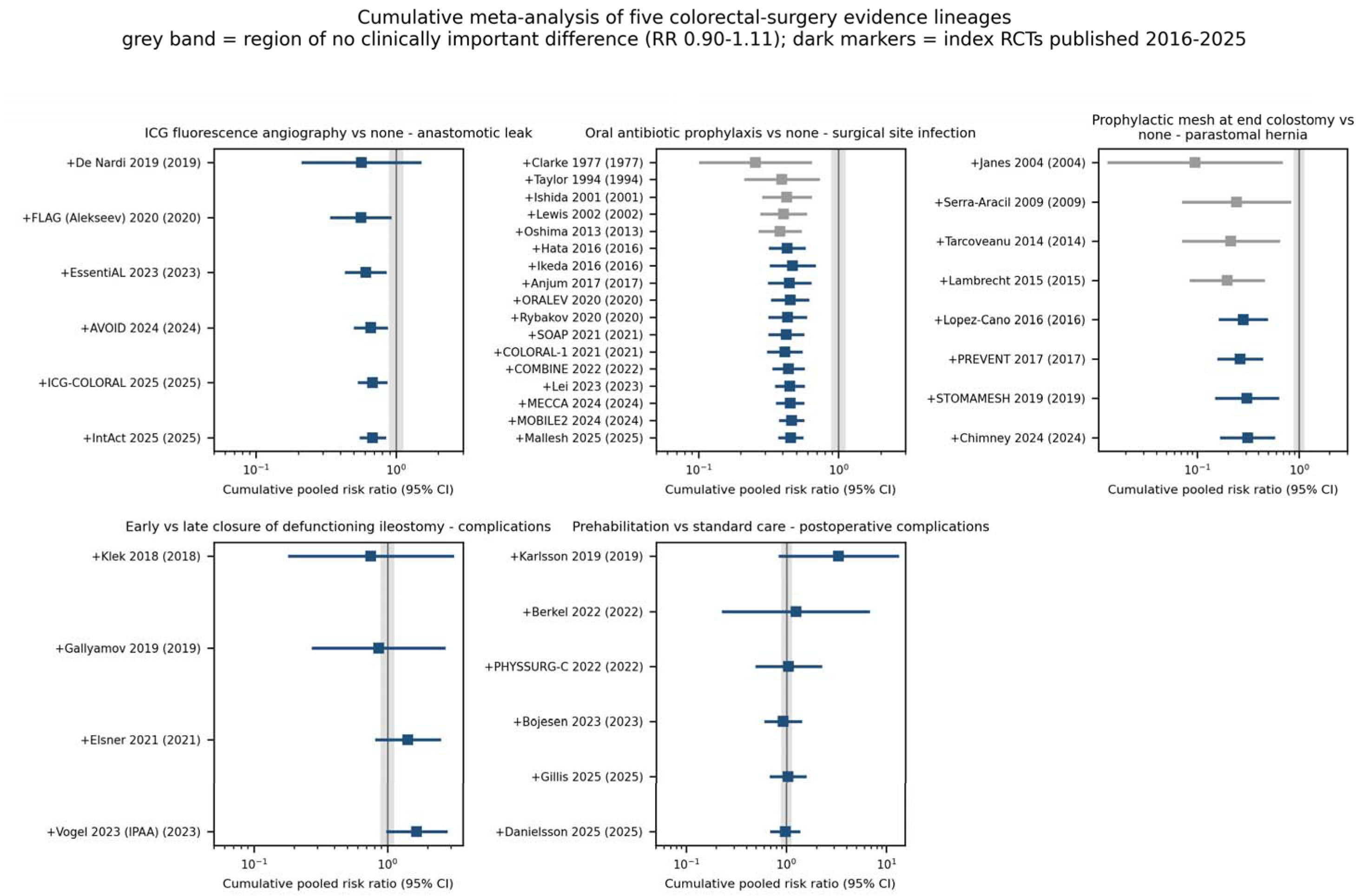
Cumulative meta-analysis of the five analyzed lineages. The shaded band is the region of no clinically important difference risk ratio 0.90 to 1.11); dark markers are trials published from 2016 through 2025, which comprise the 29 index trials and the three lineage- origin trials of that period.

**Figure 2.**
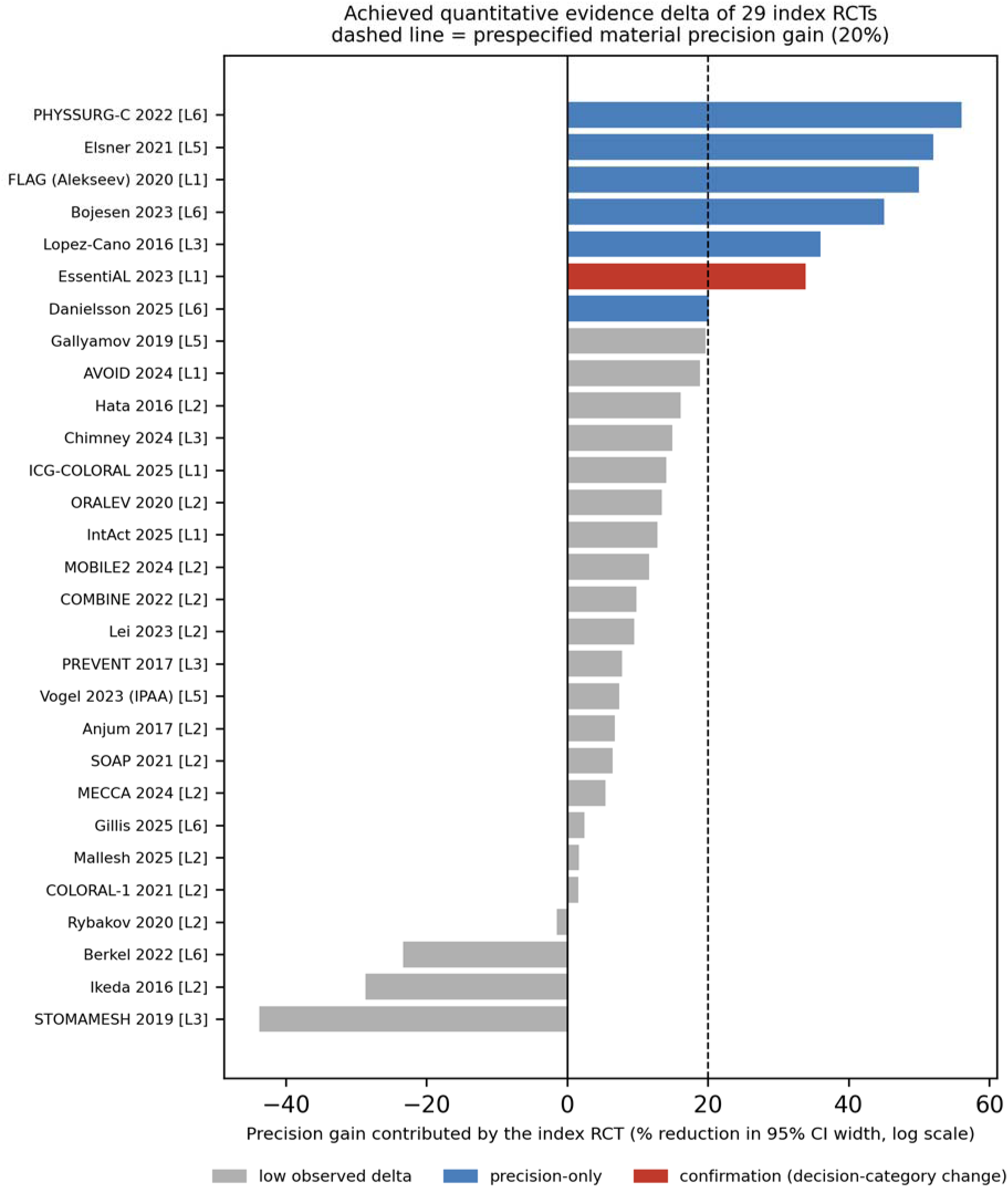
Precision gain contributed by each of the 29 index trials, ordered by magnitude and coloured by contribution class. The dashed line is the prespecified material threshold of 20%.

**Table 3.** Pre-index-result and post-index-result synthesis, decision category and evidence delta for each of the 29 index trials. Categories as in Table 2. A negative precision gain means the pooled interval widened.

| Lineage | Index trial | k<br>after | Pre-result RR (95%<br>CI) | Post-result RR (95%<br>CI) | Category | Precision<br>gain | Class |
| --- | --- | --- | --- | --- | --- | --- | --- |
| L1 | FLAG (Alekseev) 2020 | 2 | 0.56 (0.22-1.48) | 0.56 (0.35-0.90) | 4 to 4 | 50.0% | precision only |
| L1 | EssentiAL 2023 | 3 | 0.56 (0.35-0.90) | 0.61 (0.44-0.83) | 4 to 1 | 33.9% | confirmation (category change) |
| L1 | AVOID 2024 | 4 | 0.61 (0.44-0.83) | 0.66 (0.51-0.85) | 1 to 1 | 18.9% | low observed delta |
| L1 | ICG-COLORAL 2025 | 5 | 0.66 (0.51-0.85) | 0.68 (0.54-0.85) | 1 to 1 | 14.1% | low observed delta |
| L1 | IntAct 2025 | 6 | 0.68 (0.54-0.85) | 0.68 (0.56-0.83) | 1 to 1 | 12.8% | low observed delta |
| L2 | Hata 2016 | 6 | 0.38 (0.27-0.53) | 0.43 (0.33-0.57) | 1 to 1 | 16.1% | low observed delta |
| L2 | Ikeda 2016 | 7 | 0.43 (0.33-0.57) | 0.47 (0.33-0.67) | 1 to 1 | -28.7% | low observed delta |
| L2 | Anjum 2017 | 8 | 0.47 (0.33-0.67) | 0.45 (0.32-0.62) | 1 to 1 | 6.7% | low observed delta |
| L2 | ORALEV 2020 | 9 | 0.45 (0.32-0.62) | 0.45 (0.34-0.60) | 1 to 1 | 13.4% | low observed delta |
| L2 | Rybakov 2020 | 10 | 0.45 (0.34-0.60) | 0.43 (0.32-0.58) | 1 to 1 | -1.5% | low observed delta |
| L2 | SOAP 2021 | 11 | 0.43 (0.32-0.58) | 0.42 (0.32-0.56) | 1 to 1 | 6.4% | low observed delta |
| L2 | COLORAL-1 2021 | 12 | 0.42 (0.32-0.56) | 0.41 (0.32-0.54) | 1 to 1 | 1.5% | low observed delta |
| L2 | COMBINE 2022 | 13 | 0.41 (0.32-0.54) | 0.44 (0.35-0.56) | 1 to 1 | 9.9% | low observed delta |
| L2 | Lei 2023 | 14 | 0.44 (0.35-0.56) | 0.45 (0.36-0.56) | 1 to 1 | 9.6% | low observed delta |
| L2 | MECCA 2024 | 15 | 0.45 (0.36-0.56) | 0.45 (0.37-0.55) | 1 to 1 | 5.4% | low observed delta |
| L2 | MOBILE2 2024 | 16 | 0.45 (0.37-0.55) | 0.46 (0.38-0.56) | 1 to 1 | 11.7% | low observed delta |
| L2 | Mallesh 2025 | 17 | 0.46 (0.38-0.56) | 0.46 (0.38-0.55) | 1 to 1 | 1.6% | low observed delta |
| L3 | Lopez-Cano 2016 | 5 | 0.20 (0.09-0.45) | 0.28 (0.17-0.48) | 1 to 1 | 36.0% | precision only |
| L3 | PREVENT 2017 | 6 | 0.28 (0.17-0.48) | 0.26 (0.16-0.43) | 1 to 1 | 7.8% | low observed delta |
| L3 | STOMAMESH 2019 | 7 | 0.26 (0.16-0.43) | 0.31 (0.15-0.62) | 1 to 1 | -43.8% | low observed delta |
| L3 | Chimney 2024 | 8 | 0.31 (0.15-0.62) | 0.31 (0.17-0.56) | 1 to 1 | 14.9% | low observed delta |
| L5 | Gallyamov 2019 | 2 | 0.75 (0.18-3.06) | 0.86 (0.28-2.66) | 4 to 4 | 19.6% | low observed delta |
| L5 | Elsner 2021 | 3 | 0.86 (0.28-2.66) | 1.42 (0.83-2.44) | 4 to 4 | 52.1% | precision only |
| L5 | Vogel 2023 (IPAA) | 4 | 1.42 (0.83-2.44) | 1.66 (1.01-2.74) | 4 to 4 | 7.4% | low observed delta |
| L6 | Berkel 2022 | 2 | 3.30 (0.85-12.75) | 1.24 (0.23-6.56) | 4 to 4 | -23.3% | low observed delta |
| L6 | PHYSSURG-C 2022 | 3 | 1.24 (0.23-6.56) | 1.05 (0.50-2.18) | 4 to 4 | 56.1% | precision only |
| L6 | Bojesen 2023 | 4 | 1.05 (0.50-2.18) | 0.92 (0.62-1.38) | 4 to 4 | 45.1% | precision only |
| L6 | Gillis 2025 | 5 | 0.92 (0.62-1.38) | 1.03 (0.70-1.53) | 4 to 4 | 2.5% | low observed delta |
| L6 | Danielsson 2025 | 6 | 1.03 (0.70-1.53) | 0.98 (0.71-1.34) | 4 to 4 | 20.1% | precision only |

### Contextualization

Full text was retrievable for 14 of the 29 index trials, an open-access subset that is not a random sample. Among these, 8 cited a prior systematic review or meta-analysis in the introduction, 5 used one to justify the trial, its comparator, its outcome or its sample size, 8 placed the result against a prior synthesis in the discussion, and none presented an updated quantitative synthesis incorporating the new result.

### Amendments

The amendments are listed in full in the deposited amendment file; three of them affect the numbers reported here. The frozen plan specified Hartung-Knapp-Sidik-Jonkman confidence intervals as primary. On the first run these were degenerate in lineages with two or three trials, producing intervals narrower than the single-trial interval they replaced in one case and intervals spanning several orders of magnitude in another, because the t multiplier changes with the number of studies and therefore makes the pre-index-result and post-index-result intervals non-comparable. Wald intervals, which apply the same multiplier before and after, were adopted as primary and the frozen method retained as a sensitivity analysis. Under the frozen method the count would have been the same, 1 of 29, but the trial identified as changing the category would have been FLAG rather than EssentiAL. The second amendment recorded that the intracorporeal- anastomosis lineage was dropped for non-evaluability. The third amendment records that three lineages did not meet a selection criterion of the frozen plan. The fourth records that the pilot would be deposited as a preprint before any commentary citing it. The fifth records the second independent screening, the documented search and its verification search, and the independent duplicate re-extraction, which together took the syntheses from 37 trials and 27 index trials to 41 and 29 and the main-analysis proportion from 1 of 27 to 1 of 29.

## Discussion

In five colorectal-surgery evidence lineages, adding a new randomized trial to the pre-existing evidence changed the clinical decision category once in 29 trials, and under no model or threshold specification more than once. Twenty-two of 29 index trials (76%) produced neither a category change nor a material improvement in precision on the lineage outcome. These findings should not be read as a measurement of waste. They are the output of one accounting method applied to one dimension of contribution, and the same trials may have contributed safety information, applicability, or evidence for populations that this pilot did not assess.

Two results are more robust than the headline proportion. The first is that no trial changed the decision category in every specification in which it was included, and that the trial identified as the one that changed the decision was not the same trial from specification to specification. Which trial appears to change a decision depended on the interval method and on the threshold used to define clinical importance, both of which are analyst choices. Any future accounting of evidence deltas must therefore fix thresholds prospectively and report them, or the metric will be open to manipulation through the choice of specification.

The second is the reporting obstacle. More than half of the eligible randomized trials, 55%, could not enter the accounting at all. In about half of them the report does not give the number of patients with the relevant lineage outcome by treatment group; in the rest the outcome was reported only as a composite or a severity index, was not measured at all, or was reported with counts and percentages that could not be reconciled. This is not a subtle omission, and it is not new: comparing protocols with publications, Chan and colleagues found that half of efficacy outcomes per trial were reported with insufficient data for meta-analysis.[7] It defeats evidence- delta accounting, and it equally defeats any subsequent meta-analysis whose authors do not write to investigators for the missing numbers. Bibliometric measures of output register none of this. It also has a direct remedy: journals could require, for every trial, a per-arm count and denominator for each reported outcome, in the paper or in a machine-readable supplement. That single requirement would not help where the outcome was never measured, but it would recover the largest of the four groups above, which is the one in which the outcome was measured and reported without its per-arm numbers.

The lineage trajectories illustrate why a single summary statistic would mislead. In the oral- antibiotic lineage the decision category was already fixed before the study window opened, and twelve subsequent trials moved the pooled estimate from 0.38 to 0.46, a fifth of its value, while narrowing the interval by a median of 6.6% per trial. Read one way, those trials repeated a settled comparison. Read another way, they may have extended it to populations, settings and designs that the earlier trials did not cover, which the decision-category outcome cannot register and which this pilot did not assess, since the only trial characteristics extracted were the outcome definitions recorded to document imperfect harmonization. The prophylactic-mesh lineage shows the reverse pattern: the pooled estimate favored mesh from the earliest small trials, and the largest trial in that synthesis, STOMAMESH, which reported no difference in clinically judged hernia, widened the interval instead of narrowing it. A trial that increases measured uncertainty may be the most informative trial in its lineage, and any accounting that rewards precision gain alone would penalize it. This is the practical reason to keep the delta multidimensional and to report its components separately.

The pattern that cumulative meta-analysis exposed for myocardial infarction, where an effective treatment was demonstrable years before trials stopped,[4] appears here in a weaker form. In no lineage did the decision category reverse, and in four of the five it did not change at all across the decade examined. Stability of that kind is informative for trial justification, although this pilot cannot say what the evidence looked like when any of these trials was designed, since it reconstructs the evidence state immediately before each result was published and not the state at first enrollment. What it does show is that in the oral-antibiotic lineage the interval excluded the region of no clinically important difference before the study window opened and never re-entered it. Whether further trials were nevertheless warranted is a judgment about transportability, safety and local practice, and the point of an intended evidence-delta statement is to make that judgment explicit at registration rather than to leave it implicit.

The contextualization findings extend earlier work showing that systematic reviews are inconsistently used to justify new studies and rarely used to interpret them.[2,3] In the open-access subset examined here, a prior synthesis was cited more often than it was used, and although 8 of the 14 trials discussed their result against a prior synthesis, none presented an updated quantitative synthesis containing its own result. An evidence-delta statement at reporting, updating the same synthesis the trial was justified against, would close that loop with material the investigators already possess.

This pilot has substantial limitations. The lineages were chosen by the first author from fields already known to contain several trials, which may favour questions whose evidence was stable and is a source of lineage-selection bias. The sample was purposive rather than exhaustive, drawn from one database, and the archived strategies are a reconstruction that does not retrieve every record the original search found. Screening of the verification search and extraction of the pooled trials were duplicated independently, although the screening of the original interactive search is a reconstruction rather than a contemporaneous record. Every verification step that was performed, and every error it found, is recorded in the deposited audit log. The 3.4% is a feasibility estimate whose confidence interval reaches 17.2%, and it should not be quoted as a prevalence. Risk of bias, GRADE certainty, downstream use and safety and applicability deltas were not assessed. Outcome harmonization was imperfect: two trials contributed clinically relevant rather than any-grade anastomotic leak, one contributed a 90-day rather than a 30-day complication count, one lineage mixes clinically and radiologically diagnosed hernia with follow-up windows from 12 to 40 months, and one lineage includes a proctocolectomy population.

Twelve of the 41 pooled trials have counts obtained arithmetically rather than printed, and excluding them changes the main-analysis estimate to 1 of 20. The primary interval method was chosen after the first analysis run, for a documented reason, which is a post-hoc choice even though the frozen method is reported alongside. Finally, the exploratory time-locked artificial-intelligence prediction component of the parent protocol was not run, because the investigators had prior exposure to the published results and the resulting leakage is irreducible; it requires masked, prospectively generated evidence packets.

Three implications follow for research evaluation. Evidence deltas should be domain-specific and reported as components, since collapsing them into one number would recreate the problem that publication counts already cause. Thresholds must be fixed before results are known, publicly and with version history, because this pilot shows that the identity of the trial that appears to change a decision is threshold-dependent. And the accounting should be built from what investigators already hold: the same synthesis used to justify a trial, updated with its own result, would satisfy most of what is proposed here at negligible additional cost.

The next step is the full study: three databases, a peer-reviewed search strategy, prospective public registration, risk-of-bias and certainty assessment, and thresholds fixed by a panel blinded to trial results. This pilot indicates that such a study is feasible and that its rate-limiting step will be the completeness of trial reporting rather than the analysis.

## Data Availability

All study-generated data and materials are openly available in the Open Science Framework repository. The deposit contains the per-trial extraction dataset with its source-verification text; the record-level screening list for all 200 assessed records; the contextualization assessments; the frozen analysis plan and amendment log; the PubMed search strategies and yields; the verification and audit log; and the Python code with pinned package versions and archived outputs. Running the deposited code on the deposited data reproduces every statistic reported in the manuscript. Copyrighted full texts and complete PubMed records are not redistributed; each trial is identified by its PubMed identifier.

https://osf.io/mexat/

## Data availability

The deposit at https://osf.io/mexat/ contains the frozen analysis plan with its file creation date; the amendment log; the record-level screening list giving the status and exclusion reason for all 200 assessed records; the verbatim PubMed search strategies with their yields and the record of the verification search; the extraction dataset, which gives for every trial the per-arm counts, the source of each count, and the verbatim source sentence against which it was verified; the contextualization judgements with their supporting quotations; the statistical code with pinned package versions; and the verification and audit log. Running the deposited code on the deposited data reproduces every synthesis statistic reported here, including the per-lineage and per-trial values quoted in the text. The screening counts come from the screening list, the search yields from the search-strategy file, and the exclusion arithmetic and the duplicate-extraction comparison from the audit log. Full PubMed records and trial full texts are not redistributed; every trial is identified by its PMID.

## Use of artificial intelligence

The authors used a generative artificial intelligence tool (a large language model; Claude Opus 4.8, Anthropic; accessed September 2026) during preparation of this manuscript. It was used to help write and debug the Python scripts used for the statistical analysis, and to provide language editing of author-written drafts. The tool was not used to design the study, to make eligibility or study-selection judgments, or to create, alter, or interpret the original research data or results. All artificial-intelligence-assisted output was directed, reviewed, and verified by the authors, who take full responsibility for the accuracy and integrity of the content.

## Ethics approval

This meta-research study used only publicly available aggregate data from published reports and involved no individual-level participant data. Institutional review board approval and informed consent were therefore not required.

